# Genetic landscape of biofilm forming uropathogenic *E. coli* from clinical samples

**DOI:** 10.1101/2025.04.14.25325849

**Authors:** Saptarshi Roy, Deepak Jena, Mamuni Swain, Bhawna Gupta, Sunil Kumar Raghav, Surya Narayan Mishra

## Abstract

**Purpose:** Urinary tract infections (UTIs) caused by uropathogenic Escherichia coli (UPEC) are a major public health concern due to their recurrent nature and antibiotic resistance. Biofilm formation plays a crucial role in UPEC persistence, yet the genetic mechanisms underlying this process remain poorly understood. This study employs next-generation sequencing (NGS) to investigate the genomic characteristics of biofilm-forming, multidrug-resistant (MDR) UPEC isolates, with a focus on antimicrobial resistance (AMR), virulence factors, and mobile genetic elements.

**Methods:** Five biofilm-forming MDR UPEC isolates were selected for whole-genome sequencing (WGS) using the Illumina NovaSeq 6000 platform. Genome assembly and annotation were performed using SPAdes and Prokka. Multilocus sequence typing (MLST) and serotyping were conducted to determine genetic diversity, while AMR genes were identified using ResFinder. Virulence factors and biofilm-related genes were analyzed through PathogenFinder, and mobile genetic elements, including plasmids and insertion sequences, were characterized.

**Results:** Genomic analysis revealed significant diversity among isolates, with MLST identifying high-risk sequence types such as ST131, known for its strong association with MDR and virulence. AMR profiling indicated resistance to multiple antibiotics, including beta-lactams, aminoglycosides, and fluoroquinolones. All isolates harbored virulence genes associated with adhesion, immune evasion, and biofilm formation. Mobile genetic elements, particularly IncF-type plasmids and insertion sequences, were detected across isolates, suggesting a role in horizontal gene transfer of resistance traits. Biofilm-associated genes correlated with biofilm production capabilities, reinforcing their role in UPEC persistence.

**Conclusion:** This study provides critical insights into the genetic landscape of biofilm-forming UPEC, highlighting the role of mobile elements in antibiotic resistance dissemination. The findings underscore the importance of genomic surveillance and the need for novel therapeutic strategies targeting biofilm-mediated resistance to combat recurrent UTIs.

## INTRODUCTION

Urinary tract infections (UTIs) are one of the most common bacterial infections worldwide, with uropathogenic Escherichia coli (UPEC) being the primary etiological agent responsible for the majority of these infections (1). UPEC is a highly versatile pathogen capable of causing both acute and recurrent infections, often through its ability to form biofilms, which are complex communities of bacteria embedded in a self-produced extracellular matrix (2). This biofilm mode of growth provides UPEC with enhanced resistance to host immune responses, antibiotics, and environmental stresses, significantly contributing to the persistence of infections in the urinary tract (3).

The formation of biofilms by UPEC is a critical determinant of its pathogenicity and ability to persist within the host. However, the genetic basis underlying biofilm formation in UPEC remains incompletely understood. Several studies have identified a range of genes involved in biofilm formation, yet much of the genetic landscape—particularly the role of specific genetic elements and regulatory networks—remains elusive (4,5). This knowledge gap highlights the need for advanced approaches that can comprehensively assess the genetic determinants of biofilm formation in UPEC.

Next-generation sequencing (NGS) technologies, with their capacity to generate vast amounts of genomic data, offer an unprecedented opportunity to explore the genetic underpinnings of biofilm formation in UPEC (6). NGS approaches, including whole-genome sequencing (WGS) and RNA sequencing (RNA-Seq), allow for high-resolution analysis of bacterial genomes and the identification of genetic variants, gene expression profiles, and regulatory pathways that may influence biofilm formation (7). These technologies have the potential to uncover novel genetic factors involved in biofilm development, provide insights into the evolution of UPEC, and reveal therapeutic targets for disrupting biofilm-mediated UTI persistence.

This paper aims to explore the genetic landscape of biofilm-forming UPEC using NGS technologies. By leveraging these powerful tools, we seek to identify key genetic factors and regulatory mechanisms that contribute to biofilm formation in UPEC, with the ultimate goal of enhancing our understanding of UPEC pathogenesis and providing new strategies for combating recurrent UTIs. Through this work, we hope to contribute to the development of more effective treatments for UTIs that address the challenges posed by biofilm-associated bacterial resistance

## MATERIALS AND METHODS

### Bacterial Isolates and DNA Extraction

Five Escherichia coli isolates, previously identified as biofilm-forming UPEC strains with resistance to all tested antibiotics, were selected for this study. These isolates were cultured and DNA extraction was performed using a standard protocol for bacterial genomic DNA extraction. The quality and concentration of the extracted DNA were assessed using a NanoDrop spectrophotometer and electrophoresis on an agarose gel.

### Whole-Genome Sequencing

Whole-genome sequencing (WGS) was performed for all five isolates to obtain comprehensive genetic data. Libraries were constructed using the Nextera XT DNA library kit (Illumina, USA) according to the manufacturer’s instructions. Paired-end sequencing was conducted on the Novaseq 6000 sequencing platform (Illumina, USA), generating 150bp paired-end reads. Raw sequencing data were processed using the fastqc tool (v0.12.1) to assess read quality, followed by trimming of low-quality reads using the fastp tool (v0.23.2) (8)(9).

### Genome Assembly and Annotation

De novo genome assembly was performed using SPAdes v3.15.5 with the options --isolate and - k 21,31,41,51,61,71,81,91. The quality of the assembled genomes was evaluated using QUAST v5.2.0, a quality assessment tool for genome assembly. Prokka v1.13 was used to annotate the genomes, predicting coding sequences, rRNA, tRNA, and other genomic features (10).

### Identification of MLST, Serotype, Virulence, Antibiotic Resistance, and Mobile Elements

To explore the genetic diversity and pathogenicity of the UPEC isolates, we determined the multilocus sequence types (MLSTs) and serotypes of the assembled genomes using MLST v2.0 (https://cge.food.dtu.dk/services/MLST/), cgMLSTFinder v1.2 (https://cge.food.dtu.dk/services/cgMLSTFinder/), and SerotypeFinder v2.0 (https://cge.food.dtu.dk/services/SerotypeFinder/) available at the Centre for Genomics Epidemiology (CGE) server. Average nucleotide identity (ANI) values were calculated using the Integrated Prokaryotes Genome and pangenome Analysis (IPGA v1.09) to assess genome relatedness among the isolates (11).

Antimicrobial resistance (AMR) genes and point mutations responsible for resistance were identified using the Comprehensive Antibiotic Resistance Database (CARD) (12), ResFinder v4.6.0 (http://genepi.food.dtu.dk/resfinder), and VirulenceFinder v2.0 (https://cge.food.dtu.dk/services/VirulenceFinder/). The presence of virulence factors associated with biofilm formation and pathogenicity was determined using PathogeneFinder v1.1 (https://cge.food.dtu.dk/services/PathogenFinder/).

### Data Analysis and Interpretation

The genomic data obtained and analyzed to identify genetic elements related to antibiotic resistance, biofilm formation, and virulence. The isolates’ MLST profiles were compared to identify potential clonal relationships, and serotypes were determined to assess potential associations with virulence. We also evaluated the role of mobile genetic elements such as plasmids and integrons in the dissemination of antibiotic resistance and virulence factors. All plots were made using R packages.

## RESULT

### Antibiotic Resistance Profiles

Antibiotic resistance profile of five *E. coli* isolates revealed a diverse landscape of resistance and susceptibility pattern across various microbial agents (**Figure 1**). For instance, all isolates Ecoli1, Ecoli2, Ecoli3, Ecoli4, and Ecoli5 consistently exhibited resistance to cefuroxime, ceftriaxone, cefepime, and ciprofloxacin. However, fosfomycin showed susceptible to all isolates. Antibiotics like gentamicin, meropenem showed resistance to Ecoli1, Ecoli2, and Ecoli3 isolates and susceptible to Ecoli4 and Ecoli5. Four isolates except Ecoli5, showed resiastance to amoxicillin, cotrimoxazole, and piperacillin. Amikacin was found susceptible to all isolates except Ecoli2 (**Figure 1**).

**Figure 1:**
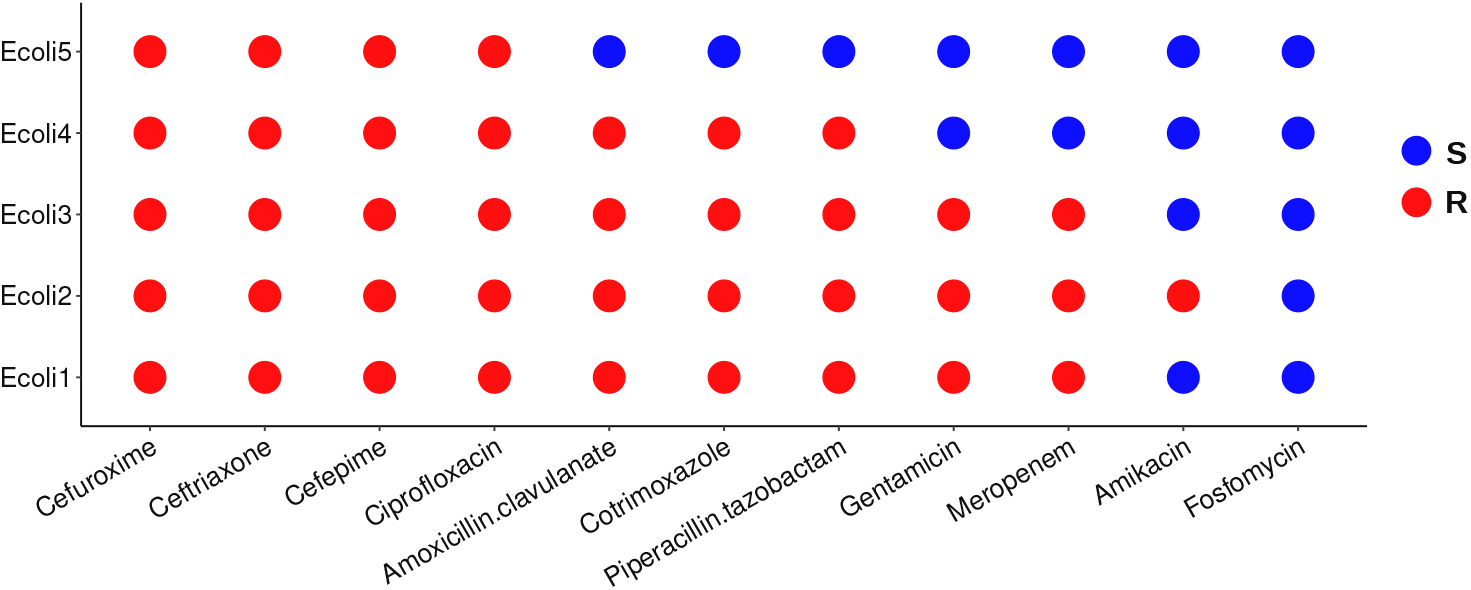
Phenotypic antimicrobial resistance profile of MDR uropathogenic *E. coli* isolates. Red color indicates resistance and blue color indicates susceptible.

### Whole genome assembly and annotation

The final assembly of *E. coli* isolates were obtained by WGS with N50 values of between 70,737 and 199,156. A total 10, 20, 9, 27, and 12 contigs representing 4,889,153; 5,366,574; 5,129,582; 6,267,315 and 4,993,494 bases were obtained from assembled sequences of *E. coli* isolates Ecoli1, Ecoli2, Ecoli3, Ecoli4, and Ecoli5 respectively, suggests that Ecoli3 has the highest-quality assembly, while Ecoli4 is more fragmented. Additionally, total 3831, 4168, 4052, 4696, 3945 genes and 5, 4, 4, 6, 5 rRNA genes were annotated from final contigs of *E. coli* isolates Ecoli1, Ecoli2, Ecoli3, Ecoli4 and Ecoli5 respectively and consistent with the expected gene content for *E. coli*, indicating the completeness of the genome assemblies (**Table 1**).

**Table 1.** Assembly Quality of five *E. coli* isolates.

| Parameter | Ecoli 1 | Ecoli 2 | Ecoli 3 | Ecoli 4 | Ecoli 5 |
| --- | --- | --- | --- | --- | --- |
| Total Length | 4,889,153 | 5,366,574 | 5,129,582 | 6,267,315 | 4,993,494 |
| Largest Contig | 325,966 | 312,350 | 403,529 | 193,832 | 358,259 |
| N50 | 171,420 | 77,897 | 199,156 | 70,737 | 137,355 |
| L50 | 10 | 20 | 9 | 27 | 12 |
| Predicted Genes | 3,831 | 4,168 | 4,052 | 4,696 | 3,945 |
| Predicted rRNA Genes | 5 | 4 | 4 | 6 | 5 |

### MLST, cgMLST, Serotype analysis

The MLST profiles determined by specific gene alleles of five *E. coli* isolates revealed common and unique sequence types (STs) in their genetic composition. Notably, Ecoli3 and Ecoli4 showed identical alleles in all seven genes resulting ST131 (**Table 2**), a globally dominant clone known for its association with extensive antimicrobial resistance (AMR) and heightened virulence. In contrast other three isolated Ecoli1, Ecoli2 and Ecoli5 have showed distinct allele type resulting ST58, ST998 and ST167 respectively, highlighting the diverse evolutionary lineages within the isolates. These results underscore the genetic heterogeneity of uropathogenic *E. coli* (UPEC) and the potential spread of high-risk clones, particularly ST131, which is notorious for causing treatment-refractory urinary tract infections (UTIs). The serotype profiles of *E. coli* isolates exhibited a variety of serotypes based on O and H antigens. Identical serotype profile was found for both Ecoli3 and Ecoli4 isolates having H5 and O16 antigen types which supports their classification as ST131, reinforcing their pathogenic potential and global prevalence. In contrast, Ecoli1, Ecoli2, and Ecoli5 presented distinct serotypes (**Table 2**).

**Table 2.** Multilocus sequence and serotyping of *E. coli* isolates.

|  |  | **Ecoli1** | **Ecoli2** | **Ecoli3** | **Ecoli4** | **Ecoli5** |
| --- | --- | --- | --- | --- | --- | --- |
| **MLST**<br>**(Multilocus**<br>**sequencing**<br>**typing)** | **adk** | 6 | 13 | 53 | 53 | 10 |
|  | **fumC** | 4 | 52 | 40 | 40 | 11 |
|  | **gyrB** | 4 | 156 | 47 | 47 | 4 |
|  | **icd** | 16 | 14 | 13 | 13 | 8 |
|  | **mdh** | 24 | 17 | 36 | 36 | 8 |
|  | **purA** | 8 | 25 | 28 | 28 | 13 |
|  | **recA** | 14 | 17 | 29 | 29 | 2 |
|  | **ST** | **58** | **998** | **131** | **131** | **167** |
| **cgMLST** |  | 145148 | 198886 | 179736 | 138639 | 191666 |
| **Serotyping** | **H type** | H30 | H6 | H5 | H5 | H9 |
|  | **O type** | O8, O179 | O2, O50 | O16 | O16 | O101 |

**Table 3.**
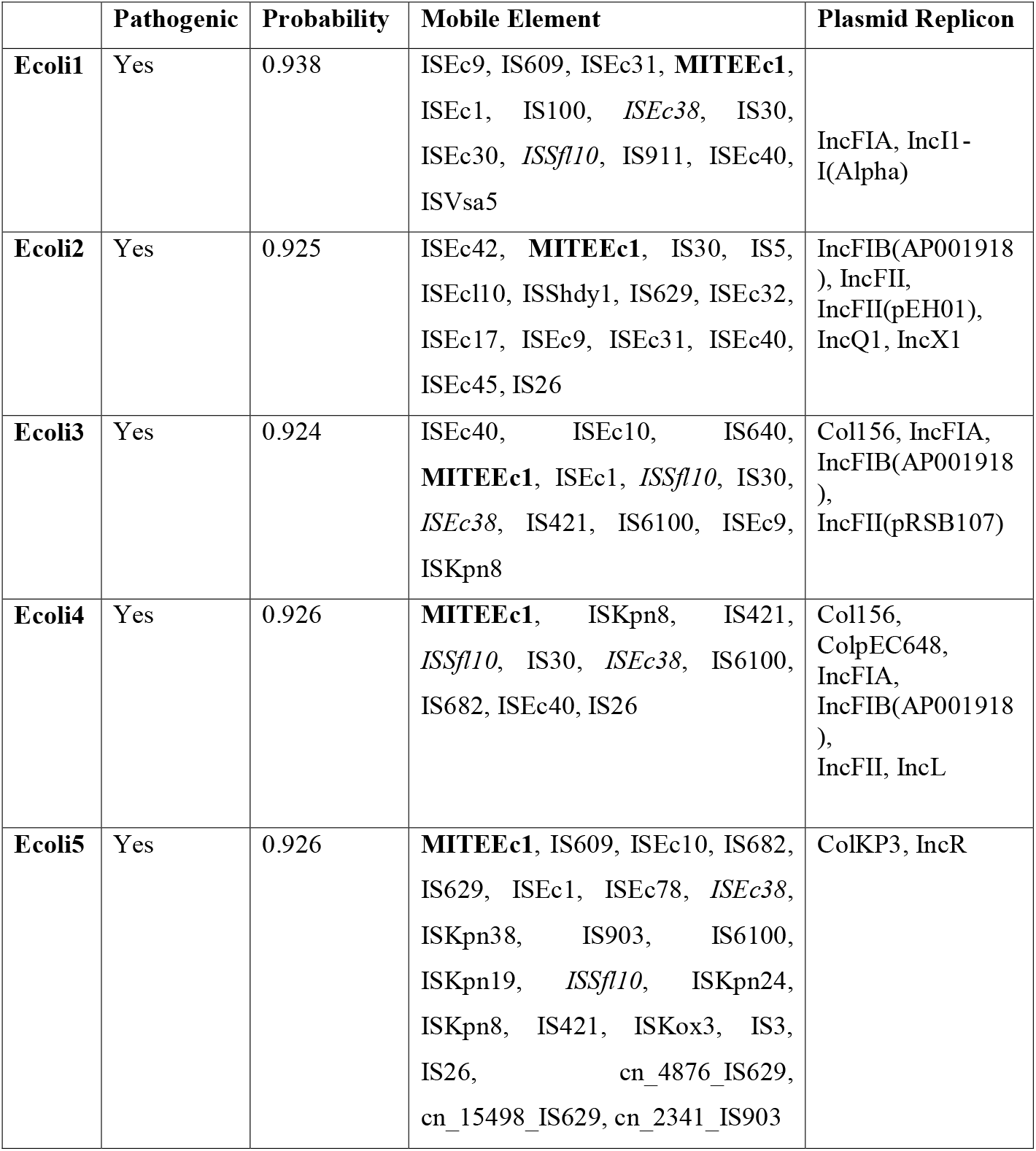
Mobile elements and plasmid replicon of E. coli chromosome. **Bold:** common in all isolates.

### Phylogenetic and ANI analysis

To assess the genomic relatedness among isolates and with other publicly available clinical *E. coli* isolates from India, phylogenomic analysis was performed using Integrated Prokaryotes Genome and pangenome Analysis (IPGA) server. Phylogenomic results indicate that Ecoli3 and Ecoli4 isolates exhibited high similarity. In contrast, Ecoli1 and Ecoli5 isolates clustered together and showed less similarity with Ecoli2 (**Figure 2A**). Further, ANI analysis were computed for all possible pairs of genome (**Figure 2B**). The *E. coli* isolates were categorized into 3 groups, with Ecoli3 and Ecoli4 forming a distinct cluster. The ANI values ranges from 96.4 to 100.00% across isolates.

**Figure 2:**
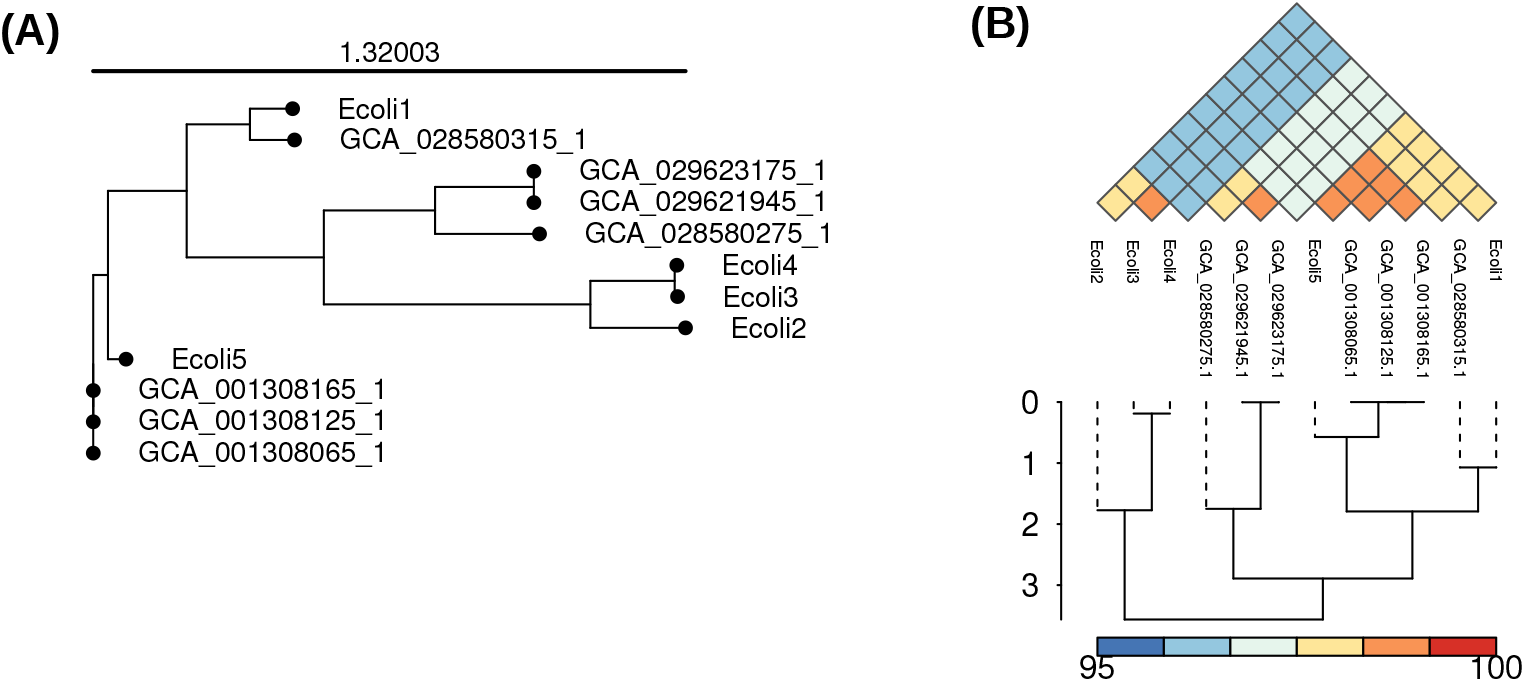
A. Phylogenetic analysis of *E. coli* isolates and publicly available clinical *E. coli* isolates. The scale bar represents genetic distance. B. ANI analysis of *E. coli* isolates and publicly available clinical *E. coli* isolates. The color indicates ANI value; ranging from 95 to 100, with color turning blue to red.

### Antimicrobial Resistance, Virulence and Biofilm producing genes in *E. coli* isolates

Antimicrobial resistance gene profiles exhibited different resistance pattern across isolates. For instance, aminocoumarin, aminoglycoside (*acrD, kdpE*), fluroquinolone (*emrA, emrB, emrR* and *mdtH*), glycopeptide (*vanG*), nitroimodazole, nucleoside, peptide and tetracycline (*emrK, emrY, mdfA*) resistance genes were universally detected in all isolates. Additionally, carbapanem, cephalosporin, diaminopyrimidine, macrolide, rifamycin, sulfonamide resistance genes varied among isolates (**Figure 3A**). Unique mutation patterns in key antibiotic resistance genes were detected among isolates, with four mutations in fluoroquinalone resistance genes (*gyrA*_D87N in Ecoli5, *gyrA*_D87Y in Ecoli3 and Ecoli4; *gyrA*_S83L in all except Ecoli1; *parC*_S80I in all except Ecoli1 and Ecoli2). A set of virulence genes (terC, yehB, yehC, yehD, nlpl, csgA, fdeC) were detected in the chromosome of all isolates (**Figure 3B**). However, Ecoli2 isolates exhibited presence of maximum number of virulence genes followed by Ecoli3 and Ecoli4 while Ecoli5 isolates showed least number of virulence genes in the chromosome. Furthermore, clustering analysis of biofilm producing genes across isolates showed Ecoli1, Ecoli2, and Ecoli4 clustered together which produces strong biofilm in the presence biofilm genes (bcsA, bcsB in Ecoli1 and Ecoli3; fimA in Ecoli3 and Ecoli4; csgA, csgB, csgC, pgaC in Ecoli4). In contrast Ecoli2 and Ecoli5 clustered together which produces weak biofilm (**Figure 3C**) (**Supplementary Table 1**).

**Figure 3:**
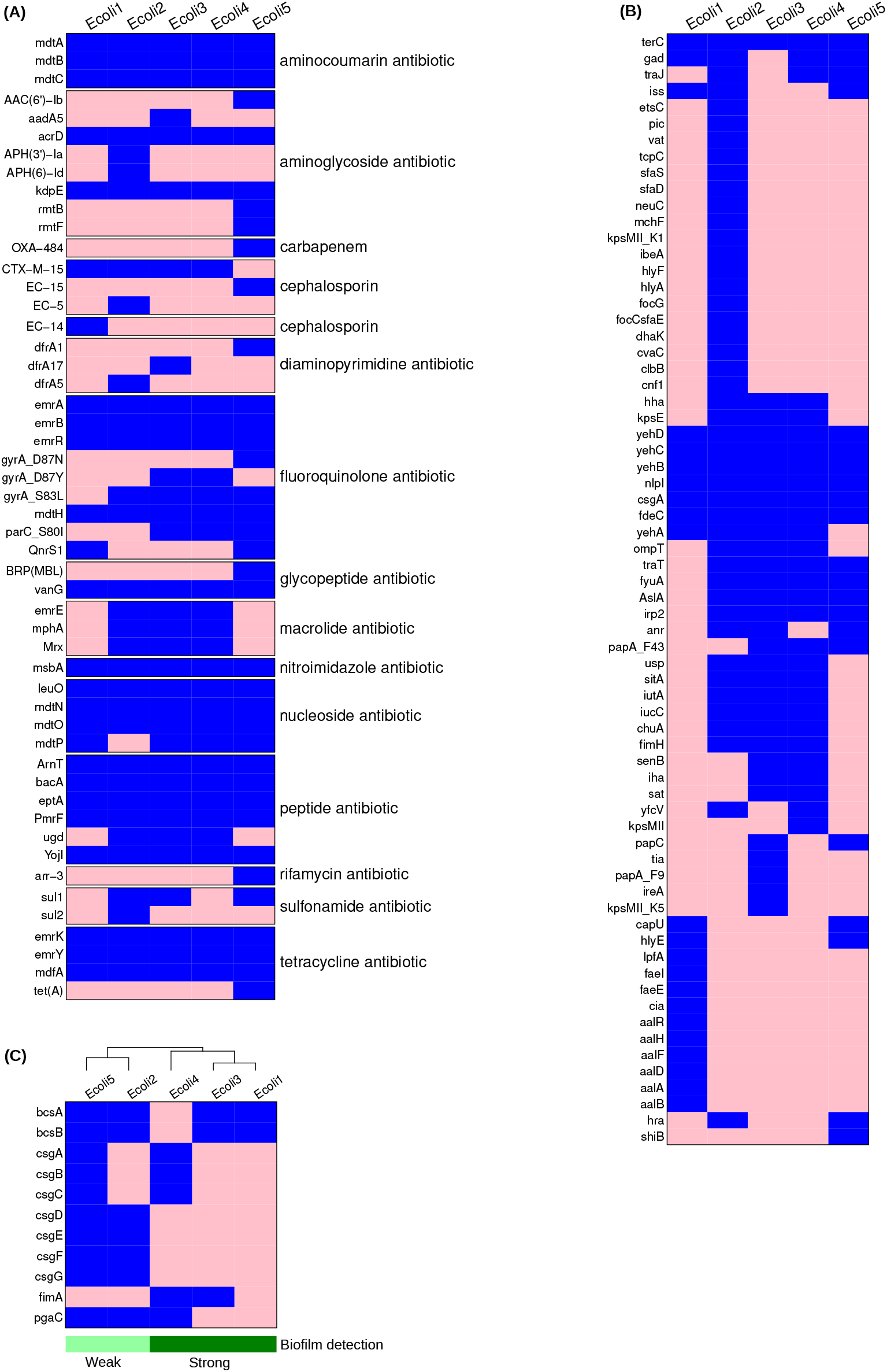
A. Antimicrobial resistance determinants in *E. coli* chromosome. Blue color indicates the presence and pink color indicates absence of antimicrobial resistance genes. B. Virulence determinants in *E. coli* chromosome. Blue color shows the presence and pink color shows absence of virulence genes. C. Biofilm determinants in *E. coli* chromosome. Blue color shows the presence and pink color shows absence of biofilm producing genes. Bar graph shows the phenotypic data related to biofilm detection.

Pathogenicity analysis revealed all five isolates are classified as pathogenic with high probabilities (0.924 – 0.938), indicating a significant potential to cause human infection, particularly urinary tract infections. Mobile element analysis showed commonalities of MITEEc1 in all isolates. Two elements ISEc38 and ISSfl10 are found present in all isolates except Ecoli2. Analysis of plasmid replicon types across all isolates revealed distinct pattern in their replication machinery. lncFIA and lncFIB replicon type were found in Ecoli1, Ecoli3 and Ecoli4 and Ecoli2, Ecoli3 and Ecoli4 respectively. Ecoli5 distinguished by featuring ColKP3 and IncR replicon type.

## DISCUSSION

The genomic analysis of the five multidrug-resistant (MDR) Uropathogenic Escherichia coli (UPEC) isolates reveals significant insights into their resistance mechanisms, biofilm-forming capabilities, and virulence potential. The detection of ST 131 in two isolates is a critical finding, as this globally disseminated clone is known for its strong association with antimicrobial resistance, particularly to fluoroquinolones and extended-spectrum beta-lactams. ST 131 is frequently implicated in urinary tract infections (UTIs) and represents a considerable therapeutic challenge in clinical practice (13).

The presence of diverse sequence types, including ST 58, ST 998, and ST 167, further highlights the genetic variability within UPEC populations. These sequence types are associated with varying levels of pathogenicity and resistance profiles, underscoring the need for genomic surveillance to track the spread of high-risk clones.

The identification of AMR genes in all isolates, coupled with the detection of plasmid replicons such as IncFIA, IncFII, and Col156, underscores the critical role of horizontal gene transfer in disseminating resistance traits (14). Plasmids not only harbor AMR genes but often carry virulence factors that enhance bacterial survival and adaptability in hostile environments. This phenomenon is particularly concerning in healthcare settings, where MDR strains can propagate rapidly under selective pressure from antibiotic use.

The high pathogenicity probabilities (0.924–0.938) obtained through computational analysis reflect the significant virulence potential of these isolates. The virulence genes identified include those responsible for adhesion, invasion, and toxin production, which are critical in the pathogenesis of UTIs. The ability to form biofilms, demonstrated by the presence of biofilm-associated genes in all isolates, further complicates the clinical management of these infections. Biofilms confer protection against both antibiotics and host immune responses, contributing to chronicity and recurrent infections, especially in catheter-associated UTIs.

## CONCLUSION

This study provides a detailed genomic profile of five MDR biofilm-forming Uropathogenic Escherichia coli (UPEC) isolates, highlighting their genetic diversity and pathogenic potential. The presence of high-risk clones such as ST 131, along with diverse AMR genes and plasmid replicons, underscores the challenges in managing infections caused by these pathogens.

The findings emphasize the need for a multifaceted approach to combat MDR UPEC infections, including:

1. Enhanced genomic surveillance to monitor the dissemination of high-risk clones and AMR genes.
2. Development of novel therapeutic strategies targeting biofilm formation and plasmid-mediated resistance.
3. Implementation of strict infection control measures to prevent the spread of MDR strains.
4. Integration of genomic tools into clinical microbiology to enable rapid identification and characterization of resistant and virulent isolates.

## Author contributions

S.R. performed the experiments, D.J. performed the data analysis and interpretation, M.S. performed the library preparation, S.R., D.J., S.K.R., and S.N.M. wrote and revised the manuscript. All authors have been approved the manuscript and contributed significantly to this work.

## Informed Consent Statement

Informed consent was obtained from all subjects involved in the study.

## Data Availability Statement

Rawdata of all five *E. coli* isolates have been deposited in the IBDC (Indian Biological Data Centre) INDA (Indian Nucleotide Data Archive) portal under the Bioproject INRP000245. INDA accessions including INS0009447, INS0009789, INS0009790, INS0009791, and INS0009874; INSDC accessions including ERS22662239, ERS22884051, ERS22886322, ERS22898190, and ERS22902670.

## REFERANCES

1. Zhou Y, Zhou Z, Zheng L, Gong Z, Li Y, Jin Y, et al. Urinary Tract Infections Caused by Uropathogenic Escherichia coli: Mechanisms of Infection and Treatment Options. Int J Mol Sci [Internet]. 2023 Jul 1 [cited 2025 Mar 12];24(13):10537. Available from: https://pmc.ncbi.nlm.nih.gov/articles/PMC10341809/

2. Whelan S, Lucey B, Finn K. Uropathogenic Escherichia coli (UPEC)-Associated Urinary Tract Infections: The Molecular Basis for Challenges to Effective Treatment. Microorganisms [Internet]. 2023 Sep 1 [cited 2025 Mar 12];11(9). Available from: https://pubmed.ncbi.nlm.nih.gov/37764013/

3. Zamani H, Salehzadeh A. Biofilm formation in uropathogenic Escherichia coli: association with adhesion factor genes. Turkish J Med Sci [Internet]. 2018 [cited 2025 Mar 12];48(1):162–7. Available from: https://pubmed.ncbi.nlm.nih.gov/29479978/

4. Alshaikh SA, El-banna T, Sonbol F, Farghali MH. Correlation between antimicrobial resistance, biofilm formation, and virulence determinants in uropathogenic Escherichia coli from Egyptian hospital. Ann Clin Microbiol Antimicrob [Internet]. 2024 Dec 1 [cited 2025 Mar 12];23(1). Available from: https://pubmed.ncbi.nlm.nih.gov/38402146/

5. Lila ASA, Rajab AAH, Abdallah MH, Rizvi SMD, Moin A, Khafagy ES, et al. Biofilm Lifestyle in Recurrent Urinary Tract Infections. Life [Internet]. 2023 Jan 1 [cited 2025 Mar 12];13(1):148. Available from: https://pmc.ncbi.nlm.nih.gov/articles/PMC9865985/

6. Satam H, Joshi K, Mangrolia U, Waghoo S, Zaidi G, Rawool S, et al. Next-Generation Sequencing Technology: Current Trends and Advancements. Biology (Basel) [Internet]. 2023 Jul 1 [cited 2025 Mar 12];12(7). Available from: https://pubmed.ncbi.nlm.nih.gov/37508427/

7. Popova L, Carabetta VJ. The use of next-generation sequencing in personalized medicine. ArXiv [Internet]. 2024 [cited 2025 Mar 12];2403.03688v1. Available from: https://pmc.ncbi.nlm.nih.gov/articles/PMC10942477/

8. Andrews S, others undefined. FastQC: a quality control tool for high throughput sequence data. 2010. 2019 [cited 2023 Aug 12];undefined-undefined. Available from: https://www.mendeley.com/catalogue/85032524-ec64-3c87-bc57-1557bf9bf08f/?utm_source=desktop&utm_medium=1.19.8&utm_campaign=open_catalog&userDocumentId=%7Bea214e00-2b4c-3ca1-9a43-fe50153662a6%7D

9. Chen S, Zhou Y, Chen Y, Gu J. fastp: an ultra-fast all-in-one FASTQ preprocessor. Bioinformatics [Internet]. 2018 Sep 1 [cited 2025 Mar 12];34(17):i884–90. Available from: 10.1093/bioinformatics/bty560

10. Seemann T. Prokka: rapid prokaryotic genome annotation. Bioinformatics [Internet]. 2014 Jul 15 [cited 2025 Mar 12];30(14):2068–9. Available from: 10.1093/bioinformatics/btu153

11. Liu D, Zhang Y, Fan G, Sun D, Zhang X, Yu Z, et al. IPGA: A handy integrated prokaryotes genome and pan-genome analysis web service. iMeta [Internet]. 2022 Dec 1 [cited 2025 Mar 12];1(4):e55. Available from: https://onlinelibrary.wiley.com/doi/full/10.1002/imt2.55

12. McArthur AG, Waglechner N, Nizam F, Yan A, Azad MA, Baylay AJ, et al. The Comprehensive Antibiotic Resistance Database. Antimicrob Agents Chemother [Internet]. 2013 Jul [cited 2025 Mar 12];57(7):3348. Available from: https://pmc.ncbi.nlm.nih.gov/articles/PMC3697360/

13. Tchesnokova V, Riddell K, Scholes D, Johnson JR, Sokurenko E V. The Uropathogenic Escherichia coli Subclone Sequence Type 131-H30 Is Responsible for Most Antibiotic Prescription Errors at an Urgent Care Clinic. Clin Infect Dis [Internet]. 2019 Feb 15 [cited 2025 Mar 20];68(5):781–7. Available from: https://pubmed.ncbi.nlm.nih.gov/29961840/

14. Flores-Oropeza MA, Ochoa SA, Cruz-Córdova A, Chavez-Tepecano R, Martìnez-Peñafiel E, Rembao-Bojórquez D, et al. Comparative genomic analysis of uropathogenic Escherichia coli strains from women with recurrent urinary tract infection. Front Microbiol. 2023 Jan 24;14:1340427.

